# What is the extent of COVID-19 vaccine hesitancy in Bangladesh? : A cross-sectional rapid national survey

**DOI:** 10.1101/2021.02.17.21251917

**Authors:** Mohammad Ali, Ahmed Hossain

## Abstract

**objectives:** To assess COVID-19 vaccine hesitancy in Bangladesh and identify population subgroups with higher odds of vaccine hesitancy.

**design:** A nationally representative cross-sectional survey was used. Univariate analysis was employed to compute vaccine hesitancy proportions and compare them across groups and multiple logistic regression analyses were performed to compute the adjusted odds ratio.

**setting:** Bangladesh

**participants:** A total of 1134 participants from the general population, aged 18 years and above.

**outcome measures:** Prevalence and predictors of vaccine hesitancy.

**results:** 32.5% of participants showed COVID-19 vaccine hesitancy. Hesitancy was high among respondents who were males, over age 60, unemployed, from low-income families, from central Bangladesh including Dhaka, living in rented houses, tobacco users, politically affiliated, participants who did not believe in the vaccine’s effectiveness for Bangladeshis and those who did not have any physical illnesses in the last year. In the multilevel logistic regression models, respondents who were transgender (AOR= 3.62), married (AOR=1.49), tobacco users (AOR=1.33), those who did not get any physical illnesses in the last year (AOR=1.49), those with political affiliations with opposition parties (AOR= 1.48), those who believed COVID-19 vaccines will not be effective for Bangladeshis (AOR= 3.20), and those who were slightly concerned (AOR = 2.87) or not concerned at all (AOR = 7.45) about themselves or a family member getting infected with COVID-19 in the next one year were significantly associated with vaccine hesitancy (p < 0.05).

**conclusions:** Given the high prevalence of COVID-19 vaccine hesitancy, it is important to promote evidence-based communication, mass media campaigns, and policy initiatives across Bangladesh to reduce vaccine hesitancy among the Bangladeshi population.

**Strengths and Limitations of the study:**

- This study is the first its kind to measure COVID-19 vaccine hesitancy in Bangladesh.
- In this study, randomly selected participants were interviewed face to face, enabling a nearly true representative sample of the Bangladeshi general population.
- This study identified a wide range of sub-groups of the general population with higher odds of COVID-19 vaccine hesitancy relating to their sociodemographic characteristics in Bangladesh; thus, providing baseline evidence for the low and middle-income and low-resourced countries worldwide.
- Traditional media and social media influence on COVID-19 vaccine hesitancy was not measured which is a major limitation of this study.

## INTRODUCTION

The first case of coronavirus disease 2019 (COVID-19) caused by severe acute respiratory syndrome coronavirus 2 (SARS-CoV-2) was detected in December 2019 in Wuhan, China. By the first week of February 2021, COVID-19 had infected over 105 million people across 223 countries or territories and caused more than 2.3 million fatalities worldwide [1]. Subsequently, COVID-19 was declared a pandemic by the WHO in March 2020 and many countries began developing COVID-19 vaccines. Two COVID-19 vaccines with 90–95% effectiveness developed by two American pharmaceutical companies were declared at the end of November 2020 [2,3]. Subsequently, many other safe and effective vaccines were also developed and announced by other countries [4–7] and by the end of 2020, 10 vaccines were approved for either full or early use in several countries including the USA, UK, and Canada [8]. Immediately after the approval, the vaccines were rolled out in the respective countries.

However, a vaccination program can be promoted or undermined by factors such as vaccine hesitancy. Vaccine hesitancy refers to delay in acceptance or refusal of vaccination despite the availability of the vaccination service [9]. In 2019, WHO declared vaccine hesitancy as one of the top ten global health threats [10]. After the COVID-19 vaccine started to rollout, besides enthusiasm, news regarding adverse effects of the vaccine experienced by a few vaccine recipients along with conspiracy theories and misinformation on social media have drawn the public’s attention worldwide [11]. Hence, puzzling news on the effectiveness of some vaccines by the media has had a negative impact on potential vaccine recipients [12,13]. Moreover, the anxiety and hesitancy is further heightened due to the accelerated pace of vaccine development [14]. Along with contemporary consequences, knowledge and awareness-related issues, vaccine hesitancy can also be determined by religious, cultural, gender, or socio-economic factors [9].

A study indicated that the vaccine willingness rate could range from 55-90% worldwide [15]. However, vaccine willingness and/or hesitancy are subject to change over time [9]. Most of the previous studies were conducted in high-income settings and well before the vaccine was made available. Very little is known about COVID-19 vaccine hesitancy in vaccination programs being run in low-income and middle-income countries’ (LMICs) population. As Bangladesh did not participate in any COVID-19 vaccine clinical trial, we hypothesized that due to the novelty of COVID-19 vaccine, there is a lack of awareness of its impact on Bangladeshis. Thus, acceptance and/or hesitancy toward the vaccine might be different compared with other available vaccines in Bangladesh.

The health, economic, and community toll of COVID-19 in Bangladesh are one of the highest among LMICs. By Mid February 2021, in Bangladesh, there were about 0.55 million laboratory-confirmed COVID-19 cases and about 10,000 died from this novel disease [16]. While the COVID-19 vaccine rollout in Bangladesh was inaugurated on 27 January 2021 targeting to immunize 138 million people [17], very little is known about COVID-19 vaccine hesitancy and/or willingness among this cohort. The government, public health officials, and advocates must be prepared to address hesitancy to reach their target and build vaccine literacy among potential recipients. Thus, our study aimed (1) to conduct a rapid national assessment of COVID-19 vaccine hesitancy in Bangladesh, and (2) to identify population subgroups with higher odds of vaccine hesitancy.

## METHODS

### Design and participants

In a cross-sectional study conducted in Bangladesh from 18 to 31 January 2021, male, female, and transgender persons aged 18 years and above were interviewed using a previously used, valid, and reliable vaccine hesitancy questionnaire [18]. To calculate sample size, a margin of error of 5%, a confidence level of 95%, a response distribution of 50% were used to target a 138 million population and secure a minimum sample size of 1067 [19,20]. Therefore, like a similar Asian study, our sample consisted of 1134 respondents [21].

### The questionnaire

The questionnaire comprised of two parts. In the first part, participants were asked questions regarding vaccine hesitancy and COVID-19 threat. First, participants were asked about the likelihood of getting a vaccine. The dependent variable and a key outcome of the study (i.e., vaccine hesitancy) were measured through the question: “If a vaccine that would prevent coronavirus infection was available, how likely is it that you would get the vaccine or shot.” The response options for this question were “Very likely,” “somewhat likely,” “not likely,” “definitely not.” Second, participants were asked two questions regarding the perceived COVID-19 threat: (1) “How likely is it that you or a family member could get infected with coronavirus in the next one year?” with response options: “very likely,” “somewhat likely,” “not likely,” and “definitely not.” (2) “How concerned are you that you or a family member could get infected with coronavirus in the next one year?” with response options: “very concerned,” “concerned,” “slightly concerned,” and “not concerned at all.”

The second part of the questionnaire comprised of a wide range of sociodemographic questions. A set of structured questions assessed participants’ gender, age, religion, marital status, education, employment status, monthly household income in Bangladeshi taka (BDT), permanent address, and region of residence in Bangladesh (north, south, and central zone including Dhaka), current residence type (Own/rented/hostel or mess), tobacco use and political affiliation. Participants were also asked about the presence of children or older people at home, whether they had any physical illnesses in the last year, whether they had a chronic disease diagnosis (hypertension, diabetes, asthma, etc.), and whether they were regular religious practitioners. These questions had to be answered using the dichotomous option (yes/no). In additions, participants were also asked two more COVID-19 vaccine-related questions: “Do you think the COVID-19 vaccine will be effective among Bangladeshis (no/yes/skeptical), and “Which developers’ vaccine would you prefer to take (American, British, Chinese, Russian, Indian, I have no idea regarding this).

### Patient and public involvement

Participants or the public were not involved in the design, or conduct, or reporting, or dissemination plans of our research.

### Data analysis

Descriptive statistics were computed to describe the demographic characteristics of the study participants. Chi-Square tests were used to compute vaccine hesitancy proportions and draw comparisons across groups. Responses were compared for various sociodemographic characteristics by dichotomizing the variable as either a positive (very likely and somewhat likely) or a negative (not likely and definitely not) attitude toward COVID-19 vaccine, indicating the extent of vaccine hesitancy. To compute adjusted odds ratios (AOR) with a 95% confidence interval, multiple logistic regression analyses were performed with vaccine hesitancy as a dependent variable and sociodemographic characteristics and perceived COVID-19 threat as predictor variables for vaccine hesitancy. To ensure the models adequately fit the data, Hosmer-Lemeshow goodness-of-fit tests were used. The significance level was set at <0.05 and SPSS version 22.0 (IBM Corp.) was used for data analyses.

## RESULTS

### Descriptive statistics analyses

Table 1 shows sociodemographic characteristics, COVID-19 threat, and vaccine hesitancy of a total of 1134 Bangladeshis who participated in this study. The mean age of the participants was 32.05 (SD ± 11.72). The majority of the study participants were: male (59.2%), aged 26-40 (40.7%), Muslim (93.2%), married (52.7%), held a bachelor’s degree (31.4%), full-time employees (28.7%), persons with a monthly household income ≥30,000 BDT (44.9%), from the central zone including Dhaka of Bangladesh (60%), living in their own house (46.3%), tobacco non-users (70.2%), those who did not get physical illnesses (57.3%) and had no political affiliation (56.5%). Regarding the question on the likelihood of getting a COVID-19 vaccine, the responses were: “very likely” (34.2%), “somewhat likely” (53.6%), “not likely” (7.3%), and “definitely not” (5.9%). In addition, **Figure 1** represents day-to-day fluctuation of vaccine hesitancy.

**Table 1.** Univariate analysis- Sociodemographic characteristics, COVID-19 threat, and vaccine hesitancy.

| Variables | Total Sample n (%) | Likelihood of getting COVID-19 Vaccine |  | p-value |
| --- | --- | --- | --- | --- |
|  |  | Not likely/definitely not n (%) | Very likely/somewhat-likely n (%) |  |
| All participants | 1134 (100%) | 369 (32.5) | 765 (67.5) | - |
| *Gender* |  |  |  | **0.003** |
| Transgender | 14 (1.2) | 9 (64.3) | 5 (35.7) |  |
| Female | 449 (39.6) | 127 (28.3) | 322 (71.7) |  |
| Male | 671 (59.2) | 233 (34.7) | 438 (65.3) |  |
| *Age group* |  |  |  | **0.009** |
| 18-25 | 442 (39.0) | 122 (27.6) | 320 (72.4) |  |
| 26-40 | 461 (40.7) | 174 (37.7) | 287 (62.3) |  |
| 41-60 | 200 (17.6) | 61 (30.5) | 139 (69.5) |  |
| ≥ 61 | 31 (2.7) | 12 (38.7) | 19 (61.3) |  |
| *Religion* |  |  |  | 0.442 |
| Muslim | 1057 (93.2) | 349 (33.0) | 708 (67.0) |  |
| Hindu | 61 (5.4) | 16 (26.2) | 45 (73.8) |  |
| Cristian and Buddhist | 16 (1.4) | 4 (25.0) | 12 (75.0) |  |
| *Marital status* |  |  |  | **0.039** |
| Unmarried | 495 (43.6) | 141 (28.5) | 353 (71.5) |  |
| Married | 598 (52.7) | 214 (35.8) | 384 (64.2) |  |
| Divorce/Widow | 42 (3.7) | 14 (33.3) | 28 (66.7) |  |
| *Children at home* |  |  |  | 0.950 |
| No | 481 (42.4) | 157 (32.6) | 324 (67.4) |  |
| Yes | 653 (57.6) | 212 (32.5) | 441 (67.5) |  |
| *Aged people at home* |  |  |  | 0.224 |
| No | 396 (34.9) | 138 (34.8) | 258 (65.2) |  |
| Yes | 738 (65.1) | 231 (31.3) | 507 (68.7) |  |
| *Education* |  |  |  | 0.268 |
| ≤ High school | 264 (23.3) | 98 (37.1) | 166 (62.9) |  |
| College education | 309 (27.2) | 92 (29.8) | 217 (70.2) |  |
| Bachelor's degree | 356 (31.4) | 111 (31.2) | 245 (68.8) |  |
| ≥ Master's degree | 205 (18.1) | 68 (33.2) | 137 (66.8) |  |
| *Employment status* |  |  |  | **0.013** |
| Full-time employee | 326 (28.7) | 109 (33.4) | 217 (66.6) |  |
| Part-time employee | 73 (6.4) | 23 (31.5) | 50 (68.5) |  |
| Business | 169 (14.9) | 66 (39.1) | 103 (60.9) |  |
| Unemployed | 88 (7.8) | 35 (39.8) | 53 (60.2) |  |
| Home maker | 171 (15.1) | 60 (35.1) | 111 (64.9) |  |
| Student | 307 (27.1) | 76 (24.8) | 231 (75.2) |  |
| *Monthly household income* |  |  |  | **0.042** |
| <15,000 | 239 (21.1) | 78 (32.6) | 161 (67.4) |  |
| 15,000-30,000 | 386 (34.0) | 108 (28.0) | 278 (72.0) |  |
| ≥ 30,000 | 509 (44.9) | 183 (36.0) | 326 (64.0) |  |
| *Family type* |  |  |  | 0.205 |
| Nuclear | 715 (63.1) | 223 (31.2) | 492 (68.8) |  |
| Joint | 419 (36.9) | 146 (34.8) | 273 (65.2) |  |
| *Permanent address* |  |  |  | 0.533 |
| Rural | 637 (56.2) | 216 (33.9) | 421 (66.1) |  |
| Urban | 411 (36.2) | 126 (30.7) | 285 (69.3) |  |
| Sub urban | 86 (7.6) | 27 (31.4) | 59 (68.6) |  |
| *Current living location* |  |  |  | **0.048** |
| Central zone | 680 (60.0) | 237 (34.9) | 443 (65.1) |  |
| North zone | 237 (20.9) | 62 (26.2) | 175 (73.8) |  |
| South zone | 217 (19.1) | 70 (32.3) | 147 (67.7) |  |
| *Current Residence type* |  |  |  | **0.042** |
| Rented | 514 (45.3) | 184 (35.8) | 330 (64.2) |  |
| Own | 525 (46.3) | 151 (28.8) | 374 (71.2) |  |
| Hostel/Mess | 95 (8.4) | 34 (35.8) | 61 (64.2) |  |
| *Regular religious practice* |  |  |  | 0.064 |
| No | 328 (28.9) | 120 (36.6) | 208 (63.4) |  |
| Yes | 806 (71.1) | 249 (30.9) | 557 (69.1) |  |
| *Tobacco user* |  |  |  | **0.037** |
| No | 796 (70.2) | 244 (30.7) | 552 (69.3) |  |
| Yes | 338 (29.8) | 125 (37.0) | 213 (63.0) |  |
| *Did you face physical illness in last one year* |  |  |  | **0.006** |
| No | 650 (57.3) | 233 (35.8) | 417 (64.2) |  |
| Yes | 484 (42.7) | 136 (28.1) | 348 (71.9) |  |
| *Morbidity* |  |  |  | 0.943 |
| No | 859 (75.7) | 280 (32.6) | 579 (67.4) |  |
| Yes | 275 (24.3) | 89 (32.4) | 186 (67.6) |  |
| *Political affiliation* |  |  |  | **0.050** |
| Ruling party | 340 (30.0) | 119 (35.0) | 221 (65.0) |  |
| Opposition | 153 (13.5) | 59 (38.6) | 94 (61.4) |  |
| Neutral | 641 (56.5) | 191 (29.8) | 450 (70.2) |  |
| *Do you think the COVID-19 vaccine will be effective among Bangladeshis* |  |  |  | **<0.001** |
| No | 108 (9.5) | 72 (66.7) | 36 (33.3) |  |
| Yes | 367 (32.4) | 43 (11.7) | 324 (88.3) |  |
| Skeptical | 659 (58.1) | 254 (38.5) | 405 (61.5) |  |
| *Which developers' vaccine would you prefer* |  |  |  | **0.001** |
| American | 435 (38.4) | 160 (36.8) | 275 (63.2) |  |
| British | 372 (32.8) | 102 (27.4) | 270 (72.6) |  |
| Chinese | 82 (7.2) | 21 (25.6) | 61 (74.4) |  |
| Russian | 64 (5.6) | 16 (25.0) | 48 (75.0) |  |
| Indian | 39 (3.4) | 8 (20.5) | 31 (79.5) |  |
| Others/no idea | 142 (12.5) | 62 (43.7) | 80 (56.3) |  |
| *Perceived likelihood of getting infected in the next 1 year* |  |  |  | **<0.001** |
| Very likely | 388 (34.2) | 141 (36.3) | 247 (63.7) |  |
| Somewhat likely | 608 (53.6) | 146 (24.0) | 462 (76.0) |  |
| Not likely | 83 (7.3) | 51 (61.4) | 32 (38.6) |  |
| Definitely not | 55 (4.9) | 31 (56.4) | 24 (43.6) |  |
| *Level of concern about getting infected in the next 1 year* |  |  |  | **<0.001** |
| Very concerned | 226 (19.9) | 30 (13.3) | 196 (86.7) |  |
| Concerned | 290 (25.6) | 53 (18.3) | 237 (81.7) |  |
| Slightly concerned | 235 (20.7) | 69 (29.4) | 166 (70.6) |  |
| Not concerned at all | 383 (33.8) | 217 (56.7) | 166 (43.3) |  |

**Figure 1.**
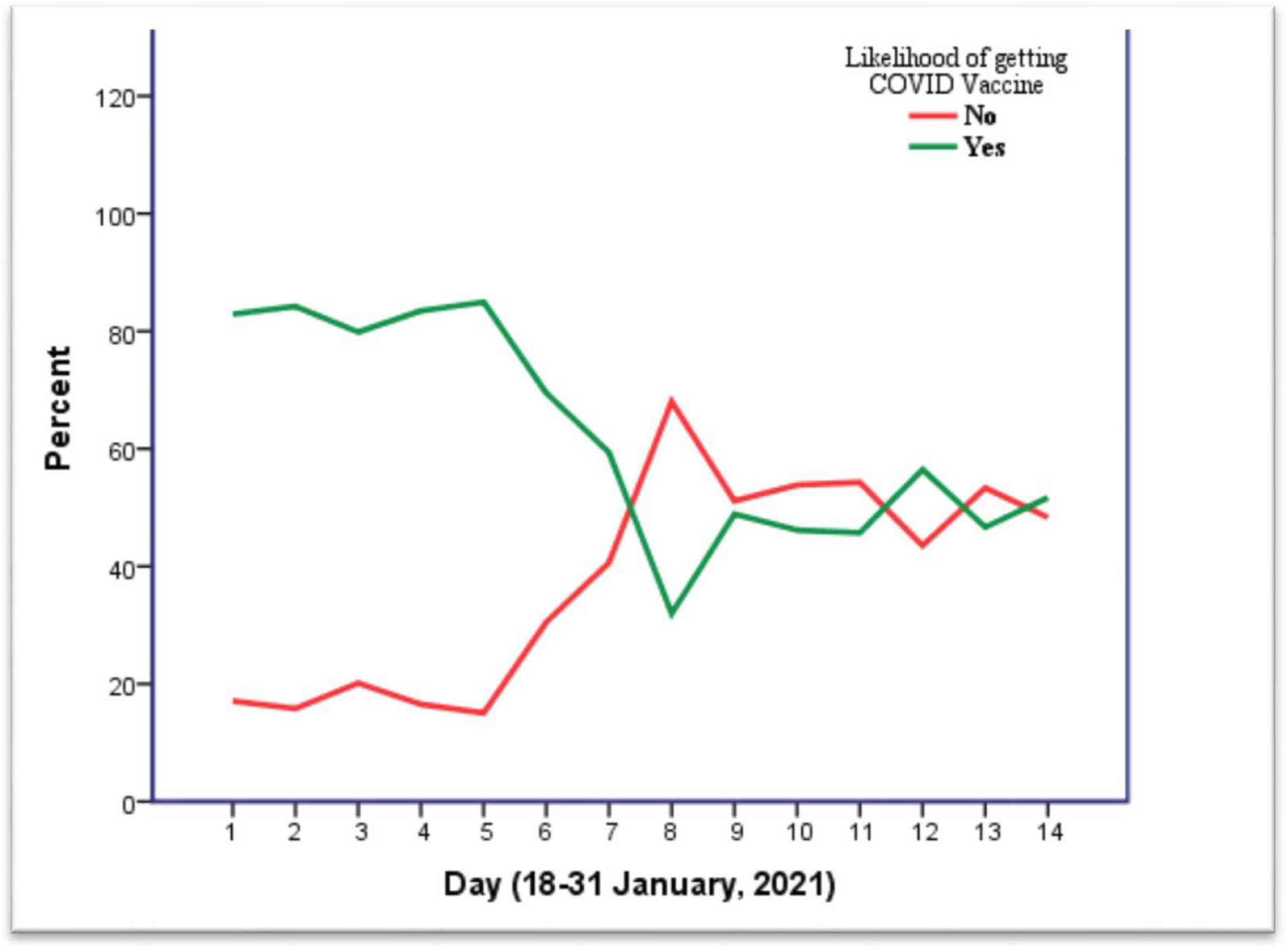
Day-to-day fluctuation of COVID-19 vaccine wiliness and/or hesitancy among Bangladeshi

### Univariate analysis

Statistically significant differences in vaccine hesitancy were found based on sociodemographic characteristics with the highest prevalence of COVID-19 vaccine hesitancy among males, persons aged over 60, businesspeople and unemployed persons, those with a monthly household income below 15 thousand BDT, living in the central zone, living in a rented house, tobacco users, those who did not face physical illness in the last year, had political affiliations with the opposition parties, did not believe in COVID-19 vaccine effectiveness among Bangladeshis, and had no knowledge on vaccine developers (**Table 1**).

Furthermore, participants who were not likely to believe that they or a family member could be infected with COVID-19 in the next one year and those who were not concerned at all about themselves or a family member getting COVID-19 infection in the next one year had the highest rates of COVID-19 vaccine hesitancy.

### Multiple logistic regression analysis

**Table 2**. presents predictors of COVID-19 vaccine hesitancy. A multiple regression analysis was conducted to examine predictors of COVID-19 vaccine hesitancy by including factors found to be significantly associated with vaccine hesitancy in the univariate analysis. In this multiple regression model, groups with statistically significantly higher odds of vaccine hesitancy were: transgender persons (AOR= 3.62, 95% CI= 1.177-11.251), married persons (AOR=1.49,CI= 1.047-2.106), tobacco users (AOR=1.33, CI= 1.018-1.745), participants who did not get physical illnesses in the last year (AOR=1.49, CI= 1.134-1.949), those with political affiliations with opposition parties (AOR= 1.48, CI= 1.025-2.134), those who did not believe in COVID-19 vaccines effectiveness for Bangladeshis (AOR= 3.20, CI= 2.079-4.925), and those who were slightly concerned (AOR = 2.87, CI= 1.744-4.721) or not concerned at all (AOR = 7.45, CI= 4.768-11.643) about themselves or a family member getting infected with COVID-19 in the next one year. Likewise, compared with participants who believed it was very likely that they or their family members could get infected with COVID-19 in the next one year, and those who thought such an occurrence would be not likely (AOR = 1.88, CI= 1.109-3.172) had significantly higher odds of vaccine hesitancy. Nonetheless, female participants (AOR= 0.70, CI= 0.537-0.928), students (AOR = 0.60, CI= 0.379-0.966) and those who preferred to take the British (AOR= 0.48, CI= 0.324-0.725), Chinese (AOR=0.44, CI= 0.245-0.807), Russian (AOR= 0.42, CI= 0.222-0.825) or Indian (AOR= 0.33, 0.143-0.774) vaccine had statistically significantly lower odds of vaccine hesitancy.

## DISCUSSION

More than one-third of the participants (32.5%) reported vaccine hesitancy in the present comprehensive national study. Analysis of daily data suggested that vaccine hesitancy varied from 18% to 72% in Bangladesh. Moreover, our study identified that predictors of COVID-19 vaccine hesitancy among Bangladeshis are gender, marital status, employment status, tobacco use, physical illness history, political affiliation, faith in vaccine effectiveness among Bangladeshis, vaccine preference, and perceived COVID-19 threat.

**Table 2.** Multiple logistic regression- predictors of vaccine hesitancy in study participants.

| Variables | Adjusted Odds Ratio | Standard Error | Confidence Interval | p-value |
| --- | --- | --- | --- | --- |
| *Gender* |  |  |  |  |
| Transgender | 3.639 | 0.576 | 1.177-11.251 | **0.025** |
| Female | 0.706 | 0.139 | 0.537-0.928 | **0.013** |
| Male | Reference |  |  |  |
| *Age group* |  |  |  |  |
| 18-25 | Reference |  |  |  |
| 26-40 | 1.208 | 0.179 | 0.851-1.715 | 0.290 |
| 41-60 | 0.808 | 0.238 | 0.508-1.285 | 0.368 |
| ≥ 61 | 1.053 | 0.434 | 0.450-2.465 | 0.905 |
| *Marital status* |  |  |  |  |
| Unmarried | Reference |  |  |  |
| Married | 1.485 | 0.175 | 1.047-2.106 | **0.027** |
| Divorce/Widow | 1.606 | 0.394 | 0.742-3.44 | 0.229 |
| *Employment Status* |  |  |  |  |
| Full-time employee | 1.006 | 0.217 | 0.657-1.539 | 0.979 |
| Part-time employee | 0.914 | 0.315 | 0.439-1.693 | 0.775 |
| Business | 1.230 | 0.227 | 0.788-1.921 | 0.362 |
| Unemployed | 1.311 | 0.284 | 0.751-2.286 | 0.341 |
| Student | 0.606 | 0.238 | 0.379-0.966 | **0.035** |
| Home maker | Reference |  |  |  |
| *Monthly household income* |  |  |  |  |
| <15,000 | Reference |  |  |  |
| 15,000-30,000 | 0.790 | 0.185 | 0.550-1.136 | 0.203 |
| ≥ 30,000 | 1.181 | 0.185 | 0.822-1.696 | 0.368 |
| *Current living location* |  |  |  |  |
| Central zone | 1.105 | 0.169 | 0.793-1.540 | 0.554 |
| North zone | 0.762 | 0.209 | 0.506-1.147 | 0.192 |
| South zone | Reference |  |  |  |
| *Current Residence type* |  |  |  |  |
| Rented | 0.962 | 0.235 | 0.607-1.527 | 0.871 |
| Own | 0.761 | 0.241 | 0.475-1.221 | 0.258 |
| Hostel/Mess | Reference |  |  |  |
| *Tobacco user* |  |  |  |  |
| No | Reference |  |  |  |
| Yes | 1.333 | 0.138 | 1.018-1.745 | **0.037** |
| *Did you face physical illness in last one year* |  |  |  |  |
| No | 1.486 | 0.138 | 1.134-1.949 | **0.004** |
| Yes | Reference |  |  |  |
| *Political affiliation* |  |  |  |  |
| Ruling party | 1.269 | 0.143 | 0.959-1.678 | 0.096 |
| Opposition | 1.479 | 0.187 | 1.025-2.134 | **0.037** |
| Neutral | Reference |  |  |  |
| *Do you think the COVID-19 vaccine will be effective among Bangladeshis* |  |  |  |  |
| No | 3.199 | 0.220 | 2.079-4.925 | **<0.001** |
| Yes | 0.212 | 0.182 | 0.149-0.303 | **<0.001** |
| Skeptical | Reference |  |  |  |
| *Which developers' vaccine would you prefer* |  |  |  |  |
| American | 0.744 | 0.197 | 0.506-1.094 | 0.133 |
| British | 0.484 | 0.205 | 0.324-0.725 | **<0.001** |
| Chinese | 0.444 | 0.304 | 0.245-0.807 | **0.008** |
| Russian | 0.428 | 0.335 | 0.222-0.825 | **0.011** |
| Indian | 0.332 | 0.431 | 0.143-0.774 | **0.011** |
| No idea | Reference |  |  |  |
| *Perceived likelihood of getting infected in the next 1 year* |  |  |  |  |
| Very likely | Reference |  |  |  |
| Somewhat likely | 0.645 | 0.161 | 0.471-0.884 | **0.006** |
| Not likely | 1.875 | 0.268 | 1.109-3.172 | **0.019** |
| Definitely not | 1.099 | 0.307 | 0.602-2.007 | 0.758 |
| *Level of concern about getting infected in the next 1 year* |  |  |  |  |
| Very concerned | Reference |  |  |  |
| Concerned | 1.609 | 0.255 | 0.977-2.649 | 0.062 |
| Slightly concerned | 2.869 | 0.254 | 1.744-4.721 | **<0.001** |
| Not concerned at all | 7.450 | 0.228 | 4.768-11.643 | **<0.001** |

This is the first study to measure COVID-19 vaccine hesitancy in Bangladesh; thus, little is known about previous hesitancy rate. However, a June 2020 global survey suggested that more than 80% of participants from China, Korea, and Singapore are very or somewhat likely to receive the COVID-19 vaccine [15]. Another study in September 2020 in Japan found 65% willingness toward the COVID-19 vaccine among participants [21]. However, a January 2021 survey in India suggested that 60% of polled Indians showed hesitancy (40% willingness) towards receiving COVID-19 vaccines [22].

We found higher vaccine hesitancy among male, older, married, and transgender participants. In the final model, women show significantly lower odds of vaccine hesitancy. In agreement with our findings, a global study observed lower odds of vaccine willingness among male participants [15]; however, women in Japan demonstrated very high vaccine hesitancy compared with men [21]. American women also showed lower willingness toward the COVID-19 vaccine [23]. Nonetheless, an early study suggested that Bangladeshi women’s better knowledge, attitude, and practice toward COVID-19 could be the reasons for their lower vaccine hesitancy [24]. An additional regional study is required to determine the gender-based difference in vaccine hesitancy. Unlike other studies, we found higher vaccine hesitancy among older people than younger. This also can be explained by an earlier study that showed lack of sufficient COVID-19-related knowledge among the older population of Bangladesh [24]. Socio-cultural and religious beliefs related to preexisting vaccine hesitancy among the older population could also be another cause of higher vaccine hesitancy. Additionally, results regarding the married population are incorporated with age; therefore, results need to be interpreted by looking at marital status and age together. Furthermore, we found statistically significant higher odds of vaccine hesitancy among the gender minority, that is, the transgender population. Previous research suggested that vaccine hesitancy is universally higher among gender minorities due to limited access and interaction with healthcare professionals, historical biomedical and healthcare-related mistrust, cost-related concerns, lack of belief in the scientific enterprise of medicine and public health, lack of awareness, and education [25].

Unemployment, an education level lower than or equal to high school, and a household monthly income less than 15,000 BDT were associated with a higher likelihood of reporting hesitancy toward COVID-19 vaccine in Bangladesh. In line with our findings, a global study also suggested that participants with lower education and income were less likely to get the COVID-19 vaccine [15]. Moreover, unemployed participants, and participants with a low level of education in the U.S. and Saudi Arabia showed higher vaccine hesitancy [23,26]. In contrast, other studies found that unemployed participants were more likely to accept the COVID-19 vaccine [20, 29]. It could be possible that in some regions, unemployed persons would like to return to work and employment and the vaccine could facilitate this return.

A unique finding of this study was that a high portion of tobacco users showed hesitancy to receive the COVID-19 vaccine. Statistically significantly high odds of vaccine hesitancy (AOR= 1.33) among tobacco users were found in the regression analysis. Universally, tobacco users (including smokers) are known to have unhealthy life practices. Further, a systematic review and meta-analysis concluded that current and previous smoking is clearly associated with severe COVID-19 outcomes [28]. Another systematic review suggested that tobacco use was significantly associated with a higher rate of mortality among COVID-19 patients [29]. So far, there have been discussions on prioritization of vaccination (e.g., for front liners). However, very little vaccination planning has been done for the most vulnerable populations who continue to remain susceptible to COVID-19 outcomes (i.e., a greater number of deaths and severe infections). Our findings would help identify these sub-groups. In contrast, we found high odds (AOR= 1.48) among those who did not have physical illnesses throughout the last year. However, previous evidence suggested that healthier persons can also be infected by COVID-19 and that the outcomes are unpredictable. Policymakers should target these subgroups when planning vaccine literacy for potential vaccine-recipients.

Interestingly, we found statically significantly higher vaccine hesitancy among politically affiliated (either affiliated with ruling parties or oppositions) participants compared with those who described themselves as neutral. However, regression analysis suggested that those who were affiliated with opposition parties had higher odds (AOR= 1.48). A systematic review and meta-analysis found a range of trust relationships with vaccine hesitancy in LMICs; for example, trust in healthcare professionals, the health system, the government, and friends and family members [30].

It is reported that the effectiveness of vaccines varies from race to race and country to country [31]. However, there was no human clinical trial of any COVID-19 vaccine in Bangladesh. In our study participants were asked whether they believed the vaccines will be effective for Bangladeshi. Those who answered “No” and remained “skeptical” showed a higher rate of vaccine hesitancy. However, this finding is similar to the findings of a study conducted in another country [32]. Finally, our study revealed very high odds of hesitancy (AOR= 7.45) among those who were not concerned about being infected by COVID-19. In support of our findings, a systematic review confirmed that people’s perceived risk of infection is one of the strongest predictors of pandemic vaccine acceptance and/or hesitancy [33].

This study result may have influenced by several limitations. Firstly, it is a cross-sectional study, and portrays a depiction of the community response at the climacteric of the study. Nonetheless, vaccine hesitancy is complex in disposition and adherence-specific, varying over time, location, and perceived behavioral nature of the community [33–35]. Secondly, social and traditional media influence are one of the major predictors of pandemic vaccine hesitancy and/or acceptance [36]. In our study, we did not examine the impact of media and this might have confounded the results. Additional research is warranted to address this issue. Despite these limitations, our study provides baseline evidence for the LMICs regarding COVID-19 vaccine hesitancy. Furthermore, our study identifies many sub-groups of the general population that must be considered during vaccine hesitancy discussions. Finally, data collected by interviewing randomly selected participants from the north, south, and central zone including Dhaka would have given a better, nearly true representative of the population of Bangladesh in the sample which would have made the study results more plausible.

## Conclusion

The present rapid community-based study on COVID-19 vaccine hesitancy in Bangladesh found that more than one-third (32.5%) of the respondents were hesitant about getting vaccinated. Differences in vaccine hesitancy were based on sociodemographic characteristics, health, and behavior of participants such as gender, age, marital status, income, employment status, tobacco use, history of illness, place of residence, and political affiliation. Further, faith in vaccine effectiveness in Bangladesh and perceived COVID-19 threat were strong predictors of COVID-19 vaccine hesitancy. Various contributing factors to vaccine hesitancy such as preexisting indecisiveness, cultural and religious views, lack of belief in the scientific enterprise of medicine and public health especially among the older population, and lower levels of awareness were identified. Further research is warranted to comprehend the complicated interplay of a variety of individual and social characteristics that influence vaccine hesitancy to ensure extensive coverage of COVID-19 vaccines. Evidence-based educational and policy-level interventions must be implemented to address these problems and promote COVID-19 immunization programs. The rates of willingness are subject to change with the suitability of the vaccines, but frequent and ambivalent effects of vaccines may reduce those rates. The uptake of COVID-19 vaccines can be increased once the factors identified in this study are properly addressed and the long-term positive effects of the vaccines are clarified.

## Data Availability

Data will be available from the corresponding author upon reasonable request.

## Acknowledgments

Both the authors acknowledge the participants for providing us the information to conduct the study.

## Data sharing statement

Data will be available from the corresponding author upon reasonable request.

## Author contributions

MA participated in study conception, design and coordination of the manuscript. MA also performed the statistical analysis and draft the manuscript. AH reviewed the manuscript and helped to draft the manuscript. Both the authors approved the final manuscript.

## Ethical consideration

Formal ethical approval was taken from the Ethical Review Committee (ERC) of Uttara Adhunik Medical College and Hospital. Prospective observational trial registration has been obtained from the World Health Organization (WHO) endorsed Clinical Trial Registry-India: CTRI/2021/01/030546. Furthermore, we conducted the study strictly following the STROBE guideline [37]. All the invited participants were required to give informed consent for participation and collection and analysis of their data.

